# Prototype abstraction predicts response to flexibility intervention in autistic youth

**DOI:** 10.64898/2026.09.01.26361990

**Authors:** Yijun Chen, Hannah Puckett, Gianna Clarot, Brylee Hawkins, Kat Sharp, Derek Alejandro Todd, Andrea Lopez, Jennifer R. Bertollo, Hannah E. Behar, Dagmar Zeithamova, Hua Xie, Alyssa Verbalis, Ashley S. VanMeter, William D. Gaillard, Lauren Kenworthy, Chandan J. Vaidya

## Abstract

Generalization is a key cognitive process that allows humans to flexibly apply prior knowledge to guide new behaviors. Difficulties with generalization and flexibility are observed across neurodevelopmental disorders, especially autism, limiting adaptive function and quality of life. Cognitive-behavioral treatment benefits some but not all autistic individuals. As treatment requires application of learned skills to everyday life, variability in generalization ability may limit intervention success in autism. While cognitive substrates of learning and generalization are well established, their potential for explaining clinical outcomes is not known. Here, we combined a category learning task with computational modelling to distinguish two learning strategies underlying generalization – prototype abstraction vs. exemplar memorization – and tested whether individual differences in these learning strategies predicted real-world intervention outcomes in autistic youth. Fifty-four participants completed the category learning task at two pre-intervention timepoints, and then completed *Unstuck and On Target:14-22* intervention targeting flexible problem solving, goal setting, and planning. We found that participants who consistently relied on prototype abstraction (N=26) were subsequently more likely to benefit from the intervention, showing improvement in parent- and self-reported flexibility. These findings identify prototype abstraction as a clinically relevant cognitive capacity that may help explain individual differences in intervention response and support the tailoring of interventions. More broadly, they demonstrate the value of linking basic cognitive mechanisms to clinical outcomes and may inform strategies to enhance the effectiveness of cognitive-behavioral interventions for youth with developmental disabilities.

## Introduction

Goal-directed flexible behavior is essential for maintaining functional independence in everyday life and adapting to changing environments. Inflexible individuals may struggle to adjust established routines, shift strategies, or respond effectively to changing demands. Difficulties with flexibility are part of broader executive functioning challenges common in autism even in the absence of intellectual disability, which include metacognitive (e.g., working memory, planning, and organization) and inhibitory domains (1). These challenges predict worse mental health (2), adaptive functioning, and quality of life (3,4), and persist even when social difficulties improve (5). Older (14-18 years) and younger (5-7 years) autistic children in particular, exhibited more difficulty with flexibility in everyday life (6). Cognitive-behavioral interventions are the first line of treatment for addressing behavioral challenges in neurodevelopmental disorders, but effects are often modest, context-bound, and vary across individuals (7–11). Understanding moderators of response variability is necessary for stratifying treatment and adapting it to non-responders for maximizing clinical outcomes (12,13). However, predicting treatment response remains an unresolved challenge (14). Cognitive-behavioral interventions require individuals not only to acquire new skills but also to generalize them beyond the treatment setting. This raises a fundamental question: Do the basic cognitive mechanisms that support learning and generalization help determine who benefits from treatment? Despite extensive study of these mechanisms in cognitive science, they have rarely been linked to clinically meaningful treatment outcomes (15).

Developmental accounts emphasize the role of abstract representation in facilitating learning and generalization in the service of flexible adaptation to the environment (16,17). One prominent form of abstract representation is prototype formation (18). Specifically, individual experiences differing in superficial details share common elements. Integrating these commonalities into a prototype that captures the central tendency of those experiences provides an efficient way to reduce memory demands and generalize to novel instances (19). Laboratory categorization tasks, coupled with computational models, have been developed to identify representations underlying learning and generalization. In a typical category learning task, participants encounter examples from contrasting categories and then categorize old and new exemplars based on the category knowledge they form. Cognitive models fitted to response patterns reveal the extent to which categorization relies on memory for specific exemplars versus an abstracted category prototype. In typically developing adults, category generalization is primarily supported by prototype representations rather than exemplar memorization (20–22). In autistic individuals, concept learning and generalization may be weaker in light of perceptual tendencies that prioritize local over global processing (23) and enhanced perceptual functioning and discrimination (24,25). However, the empirical support is mixed (reviewed in (26)). Studies using cognitive modeling in autism, although few, provide two new insights into drivers of the mixed findings. First, autistic individuals vary widely in whether they predominantly use prototype-based generalization, exemplar-based generalization, or fail to learn (27,28). Second, representational strategies may not remain stable over time across individuals (28). These observations generate testable hypotheses regarding contributions of abstract processing to variable intervention response in autism.

Here we tested whether a basic cognitive mechanism, prototype abstraction, supported response to cognitive-behavioral intervention measured with real-world outcomes in a sample of autistic youth. Participants completed assessments of category learning and flexibility at three timepoints approximately nine months apart as part of a larger longitudinal investigation of intervention outcomes (Kenworthy et al., in preparation). The first two timepoints defined a no-intervention control period, followed by *Unstuck and On Target Ages 14–22* (*Unstuck:14–22*) and the third timepoint, allowing participants to serve as their own controls. *Unstuck:14–22* is a cognitive-behavioral group intervention adapted from the established child version and targets flexibility, goal setting, and planning (29). It teaches self-regulatory strategies including integrative processing, adaptation to unexpected events, and goal-directed behavior (30–33).

Computational modelling was applied to categorization performance to infer prototype- and exemplar-based learning strategy (21,28) while flexibility was assessed with a standardized measure (34). Motivated by past results from the control period showing variability in stability of prototype-use (28), we hypothesized that youth who consistently use prototype-based category generalization prior to intervention would be better equipped cognitively to learn and generalize *Unstuck* knowledge to everyday life, thus showing greater improvements in flexibility following *Unstuck:14-22*.

## Methods and Materials

### Participants

Sixty-four adolescents with diagnosis of autism aged 14–18 years were enrolled for a longitudinal ClinicalTrials.gov enrolled study (NCT05131659) (Figure 1). Autism diagnosis was established according to DSM-5 criteria by an expert clinician, supported by a community-based diagnosis and the parent-reported Social Communication Questionnaire (SCQ), which assesses developmental history (score >7) (35). If the SCQ did not support the diagnosis or no medical diagnosis had been provided to the family, the clinician reviewed an available report describing an Autism Diagnostic Observation Schedule (ADOS) administration or, if none was available, administered the ADOS (36). In all cases, the clinician confirmed that DSM-5 criteria were met based on all available information. Medication and co-occurring conditions are reported in Supplementary Materials (SM). Inclusion criteria included Wechsler Full Scale IQ score above 80 (37), absence of neurological diagnosis (e.g., epilepsy) based on parent report, ability to participate in group therapy, and no contraindication for MRI. Out of the 64 participants who completed all assessments at T1, 54 completed the intervention and T3 assessment (see details of retention failures in SM). All participants complied with consenting procedures approved by the Institutional Review Boards at Children’s National Hospital and Georgetown University.

**Figure 1.**
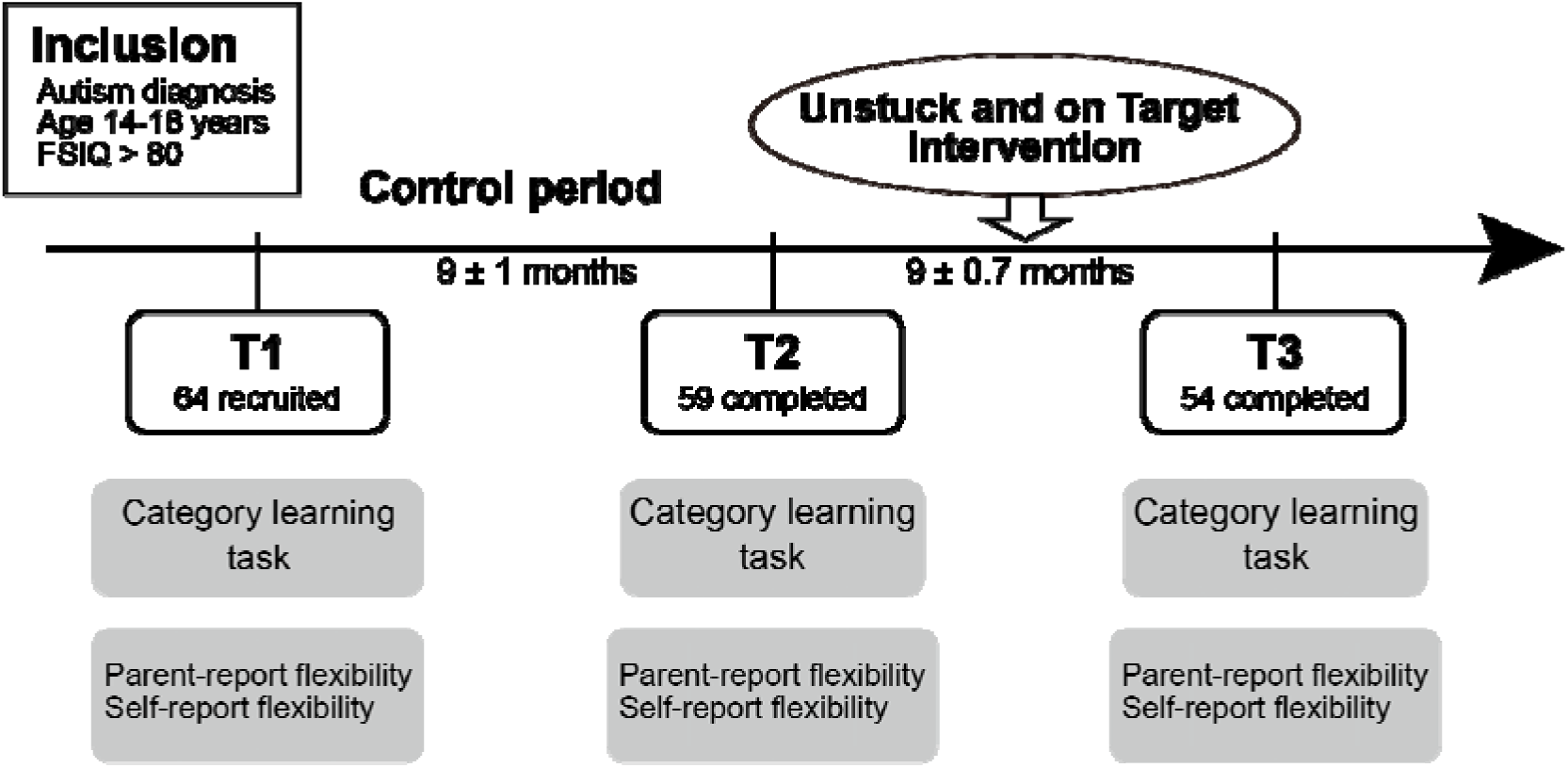
Study design and timeline. Each participant served as their own control in a longitudinal within-subject design with repeated assessments at three evenly spaced timepoints, approximately nine months apart. The interval between the first two timepoints was designated as the control period and the interval between the second and third timepoint was designated as the intervention period during which the *Unstuck:14-22* intervention was delivered in weekly group sessions. See Supplementary Materials for consort diagram.

### Study design

Participants and a parent completed assessments at the Center for Autism Spectrum Disorders at Children’s National Hospital and Georgetown University (Figure 1). At the first three timepoints, the Center visit included demographic information and parent- and self-report behavioral questionnaires, and the University visit included the category learning task and magnetic resonance imaging (MRI). An additional follow-up timepoint included behavioral assessments only. The present study reports behavioral data from the first three timepoints and category-learning data from the first two, pre-intervention timepoints. All timepoints were separated by approximately nine months. The interval from T1 to T2 constituted a no-intervention control period (range=6.4–10.4 months), followed by the *Unstuck:14-22* intervention and the interval from T2 to T3, which constituted the intervention period (range=7.5–10.2 months).

### Unstuck:14-22

*Unstuck:14-22* is a published curriculum (29) that consists of 25 one-hour group lessons. Building on earlier versions for elementary and middle schoolers (38,39), it targets flexibility, goal setting, and planning using structured teaching methods for autistic learners and individuals with executive functioning challenges, including modeling, positive reinforcement, repeated practice, visual supports, and strategy-based vocabulary (40,41). Its five units focus on understanding one’s thinking style and self-advocacy, using strategies to remain goal-directed, working flexibly with others, developing and revising plans, and consolidating learned skills.

### Behavioral assessments

#### Intervention outcome

Within a broader behavioral assessment battery, participants and a parent completed the BRIEF-2 (Behavior Rating Inventory of Executive Function-2) (34), a standardized measure of everyday executive functioning. For the present study, the eight-item Shift subscale was selected as the primary outcome because it assesses cognitive flexibility in everyday settings, including flexible problem solving, tolerance for change, attentional switching, transitions between activities, and shifting focus in daily life. Parent-reported Shift scores (parent-Shift) reflected observable flexibility difficulties in natural settings, whereas self-reported Shift scores (self-Shift) captured internal experiences of getting “stuck” and difficulties generating alternative strategies. Together, these measures provided complementary perspectives on real-world flexible behavior, with higher scores indicating greater difficulties. Raw Shift scores were used for longitudinal comparisons (analyses using T-scores yielded similar results).

#### Intervention knowledge

Participants completed an *Unstuck:14-22* knowledge questionnaire at all timepoints to assess changes in their understanding of core intervention concepts and strategies from pre- to post-intervention. It consists of 14 multiple-choice questions assessing vocabulary and concepts taught in intervention regarding flexibility, planning, self-advocacy, compromise, and goal-directed behavior. Higher scores indicated better knowledge about Unstuck.

### Category learning task

#### Materials

The category learning task was adapted from Bowman and Zeithamova (21) and employed three sets of cartoon animal stimuli, each defined by eight binary feature dimensions (Fish, Bugs, Butterflies; Figure 2A). We created three stimulus sets to maintain novelty across sessions, with a different stimulus set assigned to each session of a given participant. The order of stimulus sets across sessions was counterbalanced across participants. Within each stimulus set, one exemplar was randomly designated as the prototype for Category A, while the Category B prototype was the exemplar with all eight opposing features. Thus, the two prototypes differed on all 8 features, with their physical distance being eight features apart (Figure 2A). Category structure was defined by feature similarity to the respective prototypes. Each category included exemplars that varied by 1-away, 2-away, and 3-away from their respective prototypes. Stimuli equidistant from the prototypes (i.e., 4-away) were excluded. For the training phase, eight 2- away exemplars (four per category) were used. The generalization phase included all training stimuli, both category prototypes, as well as 24 new exemplars per category (eight 1-away, eight 2-away and eight 3-away from their respective prototypes), for a total of 58 distinct test stimuli. Within each set, participants were randomly assigned to one of four possible prototype configurations, and left/right category label placement was counterbalanced across participants.

**Figure 2.**
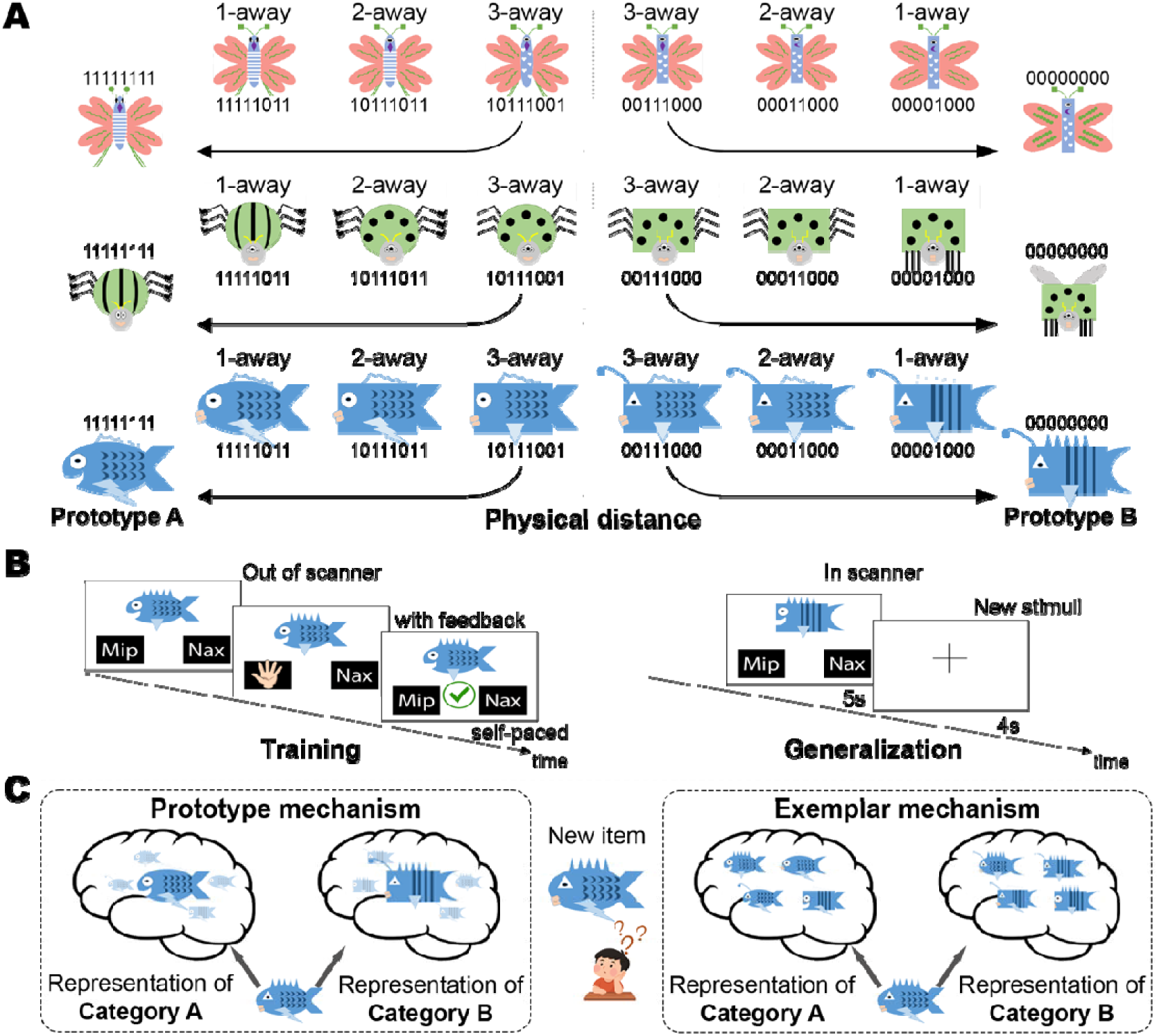
Schematic of experimental design and model fitting. (A) Representative stimuli illustrating feature distances (1-, 2-, and 3-away) from prototype A and B (0-away), for the three cartoon animal stimuli sets used in the present study. A stimulus that is 3-away from one prototype, is always 5-away from the other prototype. (B) Trial structure for the training (left) and generalization (right) phases. Training stimuli were 2-away and generalization stimuli were 0, 1, 2, and 3-away. (C) Conceptual depiction of category representations as assumed by the prototype and exemplar models.

### Training procedure

Participants viewed on-screen illustrations on a laptop and were instructed to classify the central stimuli into one of the two families by pressing a key corresponding to category labels presented on the left and right sides of the screen. They were told that at the beginning they would not know the correct answer but should guess and they would receive feedback, a correct/incorrect symbol indicating accuracy (Figure 2B left panel). Training included 240 trials, presented in five blocks of 48 trials each. Each block repeated the eight training stimuli six times in randomized order, with self-paced breaks between blocks. The prototypes were not shown. All participants proceeded to the generalization phase.

### Generalization procedure

Following training, participants completed a generalization phase in an MRI scanner across two functional MRI runs. Participants were asked to classify novel stimuli without receiving feedback, responding via two hand-held button boxes (Figure 2B right panel). Each trial included a 5-second stimulus presentation, followed by a 4-second intertrial interval during which a fixation cross was displayed. Each run included 34 trials in randomized order. These comprised eight previously shown training exemplars, two prototypes, and 24 novel stimuli (12 per category, with four at each distance: 1-, 2-, and 3-away) that differed between runs.

### Analytic plan

#### Computational models

Participants who failed to learn during training (T1: *n* = 5; T2: *n* = 6) or showed limited engagement during generalization (T1: *n* = 4; T2: *n* = 7) were not assigned a model-based categorization strategy. Failure to learn was defined as a training accuracy below .55 in the final block. Limited engagement was defined as: 1) missed >33% trials per run; 2) response bias (>75% same-button responses) with mean generalization accuracy <.55 per run; 3) task disengagement based on key-press pattern (e.g., both keys pressed, key press held through the trial or 0 reaction time).

To infer prototype- and exemplar-based representations, two similarity-based computational models (21) were fit to trial-level generalization responses in MATLAB. Both assume that learners construct internal representations and classify novel stimuli by their similarities to these representations. The models differ in how categories are represented (Figure 2C). The prototype model assumes an abstract category representation of each category (the average or ideal, defined as the most frequent feature values). In contrast, the exemplar model assumes that categories are represented by training instances (the eight training exemplars). Fit was quantified by correspondence between model-predicted and observed responses using maximum likelihood approach, separately for each model (see (28) for details). Participants were then assigned a categorization strategy based on whether they relied predominantly on prototype or exemplar representations. Strategy dominance was determined via Monte Carlo simulation (10,000 permutations), by comparing each model’s fit and the between-model difference in fit with their respective chance distributions. A strategy was classified as dominant if its observed model fit exceeded the null distribution (p <.05, one-tailed) and outperformed the competing model (difference in fit, p <.05, two-tailed). The participant was labeled as “random” if neither model exceeded chance, and as “comparable” if fits were equivalent. For subsequent analyses, participants classified as exemplar or random, as well as those not eligible for model-based strategy classification, were combined into a non-prototype category, whereas participants classified as comparable were grouped with the prototype category.

Participants who consistently exhibited prototype strategy dominance at both pre-intervention assessments were labeled as Stable. All remaining participants were categorized as Others (Figure 3). Performance accuracy for training and generalization phases for pre-intervention timepoints is presented for Stable and Other groups in SM.

**Figure 3.**
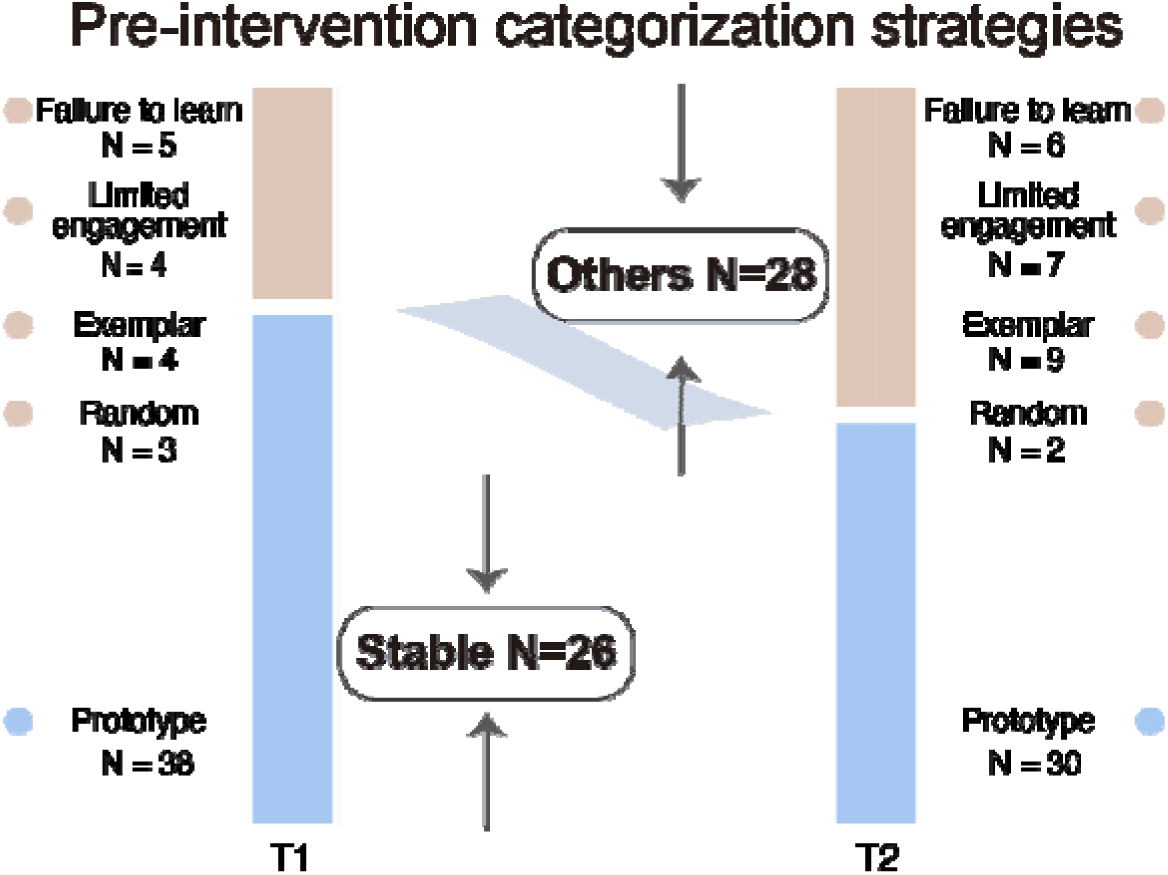
Individual differences in model-based categorization strategies at two pre-intervention timepoints. Blue represents participants classified as prototype, including one participant at T1 and one at T2 who showed comparable model-fit for prototype and exemplar strategies. Light brown represents participants classified as non-prototype, including those labeled as exemplar-dominant or random and those failing to reach training criterion or limited engagement during generalization. Participants who were prototype-dominant at both pre-intervention timepoints were designated as Stable; all others were designated as Others.

### Baseline group analyses

To characterize baseline differences, the Stable and Others groups were compared on demographic, behavioral, and clinical characteristics at T1. Independent-samples *t* tests examined demographic and background characteristics, including age, IQ, numbers of prior autism-specific and total interventions, family income, and parental education, as well as behavioral measures, including SCQ scores, flexibility, global executive functioning, and baseline *Unstuck:14-22* knowledge. Chi-square tests examined categorical characteristics, including sex assigned at birth, self-reported gender (M, F, and non-binary), medication categories, and co-occurring conditions.

### Main analyses

Hierarchical linear modeling (HLM) implemented in MATLAB using the “fitlme” function was used to account for the hierarchical structure of the data. To evaluate whether the Stable and Others groups exhibit different longitudinal trajectories of Shift scores across the intervention, we tested the *timepoint × group* interaction. Significant interactions were followed by post hoc analyses within each group. The dependent variables were raw parent- and self-Shift scores. All three timepoints were included in a single model, with timepoint dummy-coded using T2 as the reference. This coding yielded two timepoint contrasts corresponding to the control period (T1 to T2) and the intervention period (T2 to T3). Accordingly, two *timepoint × group* interaction terms tested whether the groups differed in change during the control and intervention periods, respectively. Age at baseline, sex assigned at birth (M, F), IQ and SCQ score at baseline were included as covariates. Outcome measure for timepoint j, subject i was modeled as:

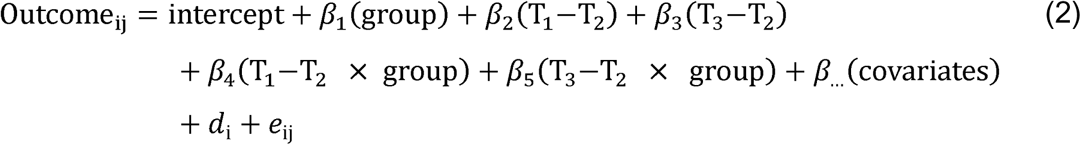

Here, the intercept and β terms are fixed effects, d_i_ is the subject-level random effect, and e_ij_ is the residual. The intercept and slope of timepoint were allowed to vary across individuals.

### Control analyses

To examine the robustness of the *timepoint × group* interaction, two control analyses were conducted. First, as the Others group was heterogeneous, comprising participants who exhibit no consistent strategy, failed to learn, or showed limited engagement, we repeated the analyses to compare the Stable group with only those without consistent learning strategy, by excluding participants who failed to learn or showed limited engagement. Second, we repeated the main analysis using two alternative single-timepoint definitions of the Stable group, based on prototype dominance either at T1 or T2.

## Results

### Stable vs. Others prototype representation

We first classified learning strategy at the two pre-intervention timepoints. Most participants were prototype-dominant at each assessment: 70% (*n* = 38) at T1 and 56% (*n* = 30) at T2. Of those classified as prototype-dominant at T1, 68% (*n* = 26) remained so at T2 (Figure 3). Stable prototype representation was defined as prototype dominance at both pre-intervention assessments, yielding 26 participants in the Stable group and 28 in the Others group (Table 1).

**Table 1.**
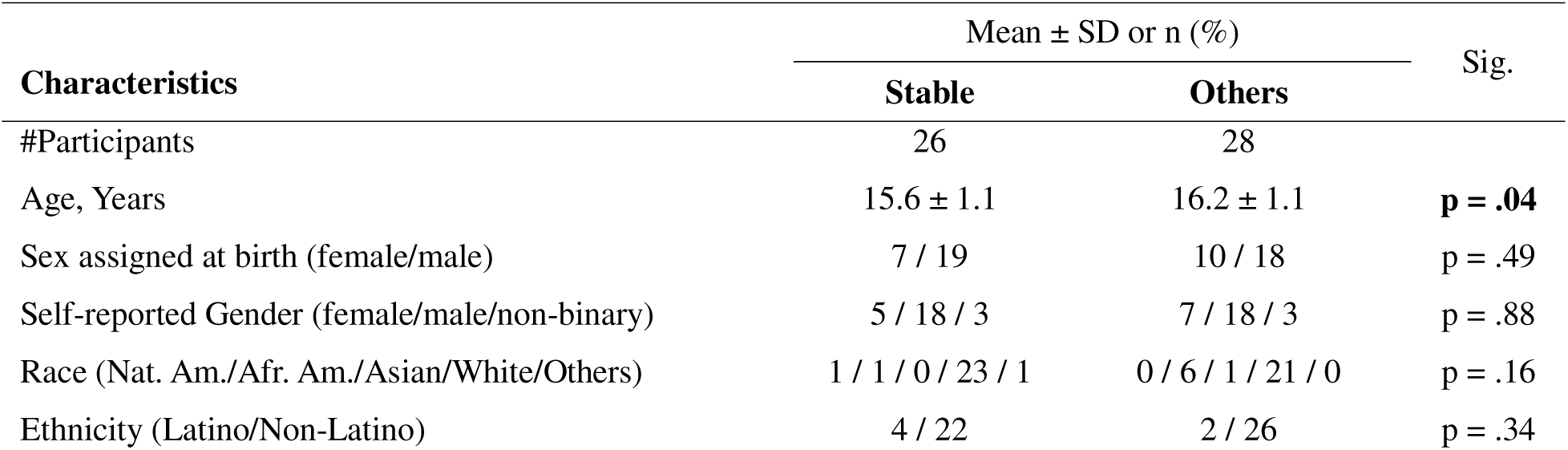
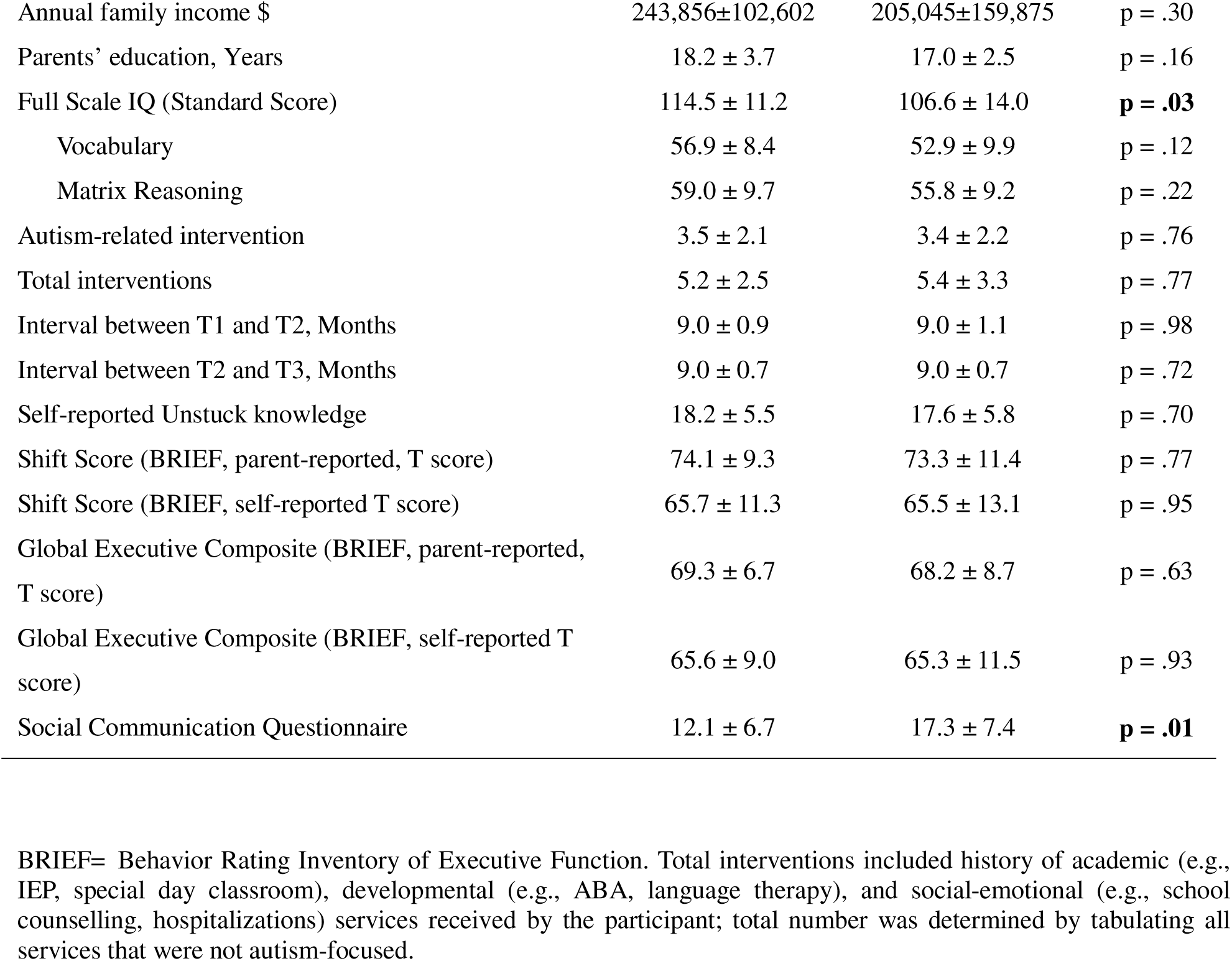
Demographic and baseline (T1) characteristics for Stable and Others groups.

| Characteristics | Mean ± SD or n (%) | | Sig. |
| --- | --- | --- | --- |
|  | Stable | Others |  |
| #Participants | 26 | 28 |  |
| Age, Years | 15.6 ± 1.1 | 16.2 ± 1.1 | **p = .04** |
| Sex assigned at birth (female/male) | 7 / 19 | 10 / 18 | p = .49 |
| Self-reported Gender (female/male/non-binary) | 5 / 18 / 3 | 7 / 18 / 3 | p = .88 |
| Race (Nat. Am./Afr. Am./Asian/White/Others) | 1 / 1 / 0 / 23 / 1 | 0 / 6 / 1 / 21 / 0 | p = .16 |
| Ethnicity (Latino/Non-Latino) | 4 / 22 | 2 / 26 | p = .34 |
| Annual family income \$ | 243,856±102,602 | 205,045±159,875 | p = .30 |
| Parents' education, Years | 18.2 ± 3.7 | 17.0 ± 2.5 | p = .16 |
| Full Scale IQ (Standard Score) | 114.5 ± 11.2 | 106.6 ± 14.0 | **p = .03** |
| Vocabulary | 56.9 ± 8.4 | 52.9 ± 9.9 | p = .12 |
| Matrix Reasoning | 59.0 ± 9.7 | 55.8 ± 9.2 | p = .22 |
| Autism-related intervention | 3.5 ± 2.1 | 3.4 ± 2.2 | p = .76 |
| Total interventions | 5.2 ± 2.5 | 5.4 ± 3.3 | p = .77 |
| Interval between T1 and T2, Months | 9.0 ± 0.9 | 9.0 ± 1.1 | p = .98 |
| Interval between T2 and T3, Months | 9.0 ± 0.7 | 9.0 ± 0.7 | p = .72 |
| Self-reported Unstuck knowledge | 18.2 ± 5.5 | 17.6 ± 5.8 | p = .70 |
| Shift Score (BRIEF, parent-reported, T score) | 74.1 ± 9.3 | 73.3 ± 11.4 | p = .77 |
| Shift Score (BRIEF, self-reported T score) | 65.7 ± 11.3 | 65.5 ± 13.1 | p = .95 |
| Global Executive Composite (BRIEF, parent-reported, T score) | 69.3 ± 6.7 | 68.2 ± 8.7 | p = .63 |
| Global Executive Composite (BRIEF, self-reported T score) | 65.6 ± 9.0 | 65.3 ± 11.5 | p = .93 |
| Social Communication Questionnaire | 12.1 ± 6.7 | 17.3 ± 7.4 | **p = .01** |

BRIEF= Behavior Rating Inventory of Executive Function. Total interventions included history of academic (e.g., IEP, special day classroom), developmental (e.g., ABA, language therapy), and social-emotional (e.g., school counselling, hospitalizations) services received by the participant; total number was determined by tabulating all services that were not autism-focused.

### Baseline group differences in demographic and behavioral characteristics

At baseline, the Stable group was younger (*t*(52)=2.1, *p*=.04), and had higher IQ (*t*(52)=2.3, *p*=.03). The groups did not differ in other demographic or background characteristics, including sex, gender, family income, parental education, or prior interventions (*p*s≥.16; Table 1). On behavioral measures, the Stable group had lower autistic traits across developmental history, as measured by the SCQ (*t*(52)=2.7, *p*=.01), but did not differ in flexibility, global executive functioning, or self-reported Unstuck knowledge (all *p*s≥.63; Table 1). The groups also did not differ in number of co-occurring conditions, or medication status (all *p*s≥.17; Table S1).

### Different outcome patterns between Stable and Others groups

Of main interest was the degree to which the ability to form abstract representations predicts intervention outcomes. If abstract prototype formation supports generalization of intervention treatments to everyday life, the Stable group should show greater intervention-related improvement than the Others group. In the HLMs, the main effect of group was not significant for either parent- or self-Shift (*p*s≥.60), indicating that overall, across timepoints, groups did not differ in flexibility. We then tested the *timepoint × group* interaction from T2 to T3 to determine whether the groups differed in intervention-related changes in flexible functioning. The interactions were significant for both the parent- and self-Shift (parent-Shift: *t*(152)=2.2, *p*=.03; self-Shift: *t*(152)=2.3, *p*=.02; Figure 4), indicating differential intervention response between groups. Post-hoc tests showed that the Stable group improved significantly after intervention (parent-Shift: *t*(71)=-3.7, *p*<.001; self-Shift: *t*(72)=-2.8, *p*=.007), while the Others group did not differ across timepoints (parent-Shift: *p*=.23; self-Shift: *p*=.57). To determine whether the group differences were specific to the intervention period, we also examined the *timepoint × group* interaction from T1 to T2, when any change would reflect the passage of time rather than intervention. Neither interaction was significant (parent-Shift: *p*=.73; self-Shift: *p*=.14). Together, these results suggest that the Stable group exhibited greater improvements in flexible functioning specifically during the intervention period, rather than nonspecific changes over time.

**Figure 4.**
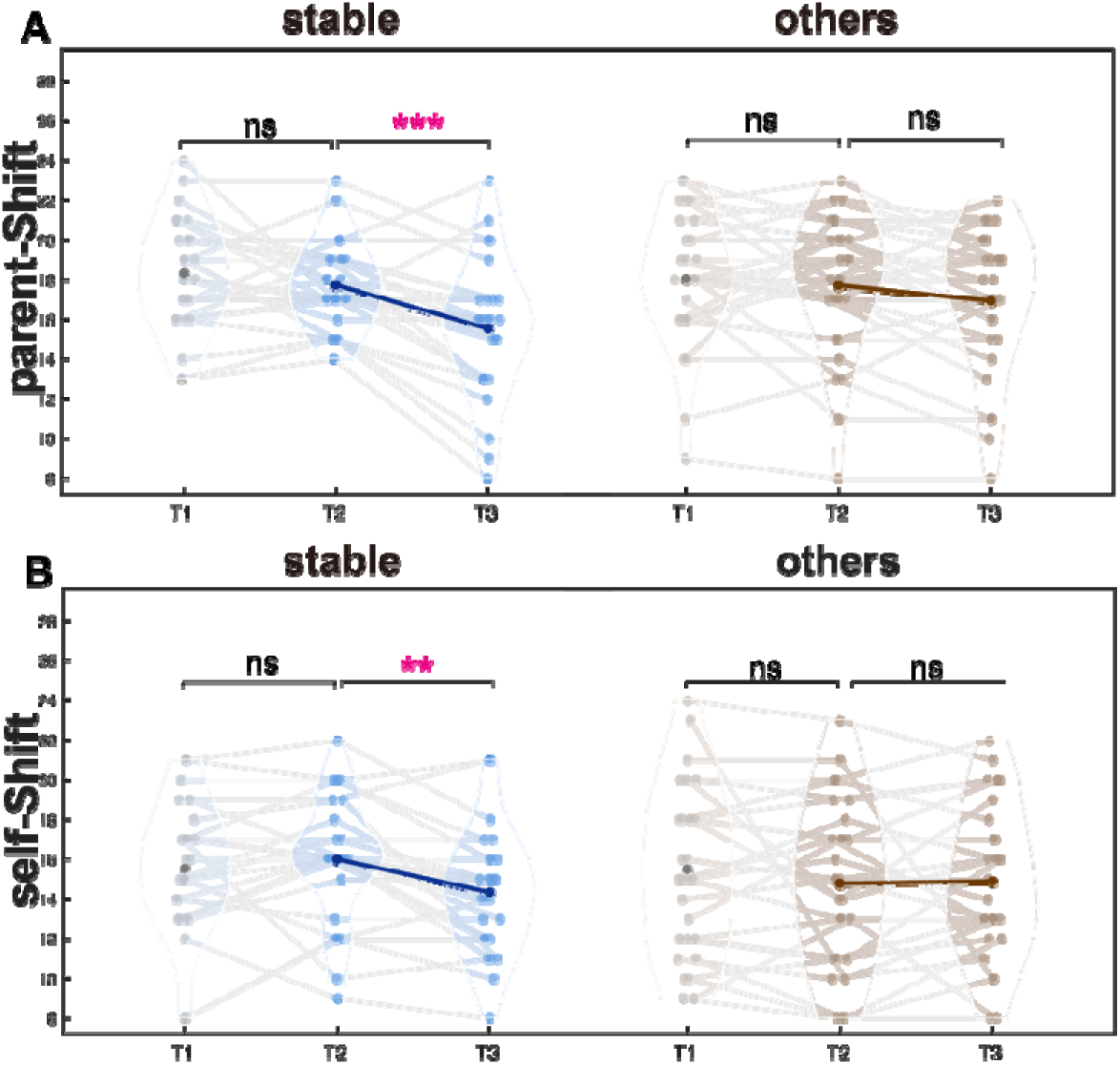
Flexibility improvements following intervention in the Stable but not Other group. (A) Parent-reported flexibility measure, (B) Self-reported flexibility measure at T1, T2, and T3 for the Stable (*n* = 26) and Others (*n* = 28) groups. Blue indicates the Stable group, and brown indicates the Others group. Violin plots illustrate the distribution of scores at each timepoint, with individual data points overlaid. Gray connecting lines represent within-subject trajectories across the three timepoint. Dark and bold lines depict group mean changes during the intervention. _∗∗_=*p* < .01; _∗∗∗_=*p* < .001; ns=not significant. Lower value = greater flexibility.

In light of the observed intervention-related change in flexibility selectively in the Stable group, we tested whether knowledge about the intervention also differed similarly between groups using the same HLM model. The *timepoint × group* interactions were significant (*t*(152)=-2.4, *p*=.02, Figure 5), indicating differential intervention knowledge between groups. Post-hoc tests showed that the Stable group demonstrated significantly higher *Unstuck* knowledge after intervention (*t*(71)=5.2, *p*<.001), while it did not differ across timepoints in the Others group (*p*=.17). *Timepoint x group* interactions were not significant for the control period, T1 to T2 (*p*=.93) indicating that change in intervention knowledge was specific to the intervention period and did not extend simply to the passage of time.

**Figure 5.**
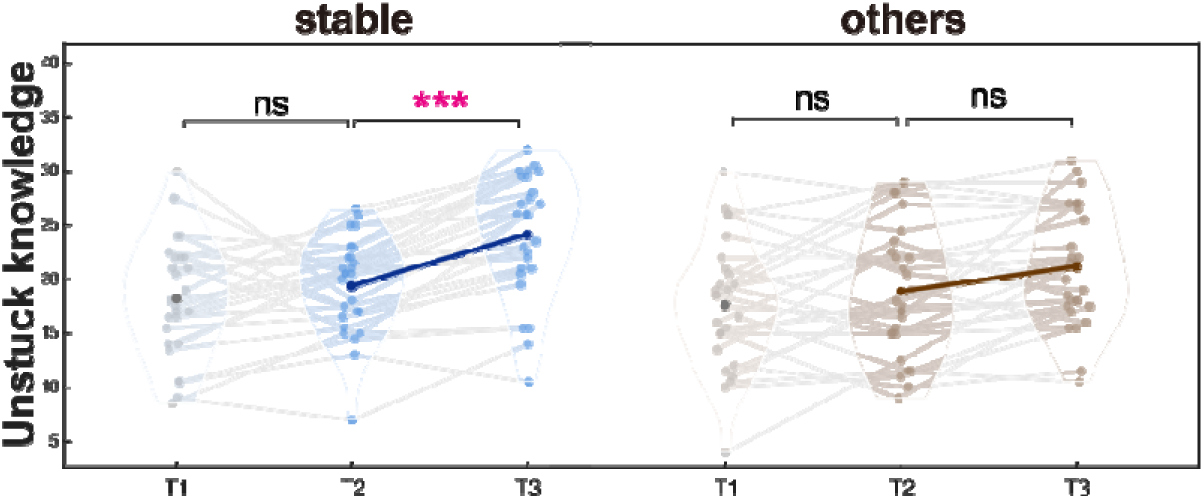
Greater Unstuck 14–22 knowledge gains in the Stable group. Self-reported knowledge about Unstuck concepts and strategies measured at T1, T2, and T3 for the Stable (*n* = 26) and Others (*n* = 28) groups. Blue indicates the Stable group, and brown indicates the Others group. Violin plots illustrate the distribution of scores at each timepoint, with individual data points overlaid. Gray connecting lines represent within-subject trajectories across the three timepoint. Dark and bold lines depict group mean changes during the intervention. _∗∗∗_=*p* < .001; ns=not significant.

### Control analyses for flexibility improvements

To examine the robustness of the *timepoint × group* interactions of flexibility scores, we conducted two control analyses. First, after excluding 16 participants who failed to learn and/or showed limited engagement from the Others group, the interaction remained significant for parent-report (*t*(105)=2.4, *p*=.02), and marginally for self-report (*p*=.11). Second, we repeated the analyses using prototype dominance at a single pre-intervention timepoint to define Stable group. When group assignment was based on T1, the interaction was significant for parent-report flexibility (*t*(153)=2.1, *p*=.04), but not for self-report (*p*=.27). When assignment was based on T2, the opposite pattern emerged, the interaction was significant for self-report (*t*(153)=2.2, *p*=.03), but not for parent-report (*p*=.37). Thus, single-timepoint classifications produced inconsistent findings across outcomes, supporting the use of repeated assessments to identify reliable prototype representation.

## Discussion

The present study tested the potential of a cognitive capacity for reliable use of abstract processing in concept generalization, to predict responsiveness to a cognitive-behavioral flexibility intervention in autistic youth. This work was motivated by individual differences observed in a longitudinal study examining pre-intervention representational strategies during category generalization (28). Approximately half of the sample reliably utilized prototype information for categorization across two measurements obtained nine months apart. Consistent with our hypothesis, these individuals derived greater benefits from subsequent participation in *Unstuck:14-22*, as reflected in improved flexibility in everyday life reported by parents and youth themselves. In contrast, individuals who did not exhibit consistent prototype abstraction did not show reliable flexibility improvements following intervention. These findings provide empirical support for the theorized role of category abstraction and generalization processes in adaptive learning and transfer (19), and advance translational efforts by explaining one source of heterogeneity in intervention response across autistic individuals.

Our findings highlight the contribution of abstract concept representation to variability in cognitive-behavioral intervention response in autism. Although the majority of autistic youth showed prototype-dominant generalization at each assessment, only about half maintained this pattern across two pre-intervention assessments. Thus, despite demonstrating a capacity for abstract representation, many did not draw on it consistently. Reliable prototype use may, therefore, reflect a trait-like cognitive strategy for abstraction and generalization. This cognitive trait could be particularly advantageous for *Unstuck:14-22*, a top-down, strategy-based intervention that relies on the acquisition and transfer of vocabulary scripts (e.g., “big picture” thinking for integration, “plan B” for coping with unexpected events, “eyes on the prize” for goal-setting) across contexts. Individuals who consistently relied on abstract concept representations may have been better able to internalize and generalize *Unstuck* scripts across situations. This interpretation is supported by their greater gains in *Unstuck* knowledge during the intervention (Figure 5), which may have aided application of *Unstuck* skills to everyday life. Importantly, these gains were not explained by baseline flexibility or broader executive functioning, as groups did not differ in either domain. The groups also did not differ in composition of sex, gender, race, socioeconomic characteristics, intervention history, or time between assessments. Medication status and concurrent conditions also did not differ between groups. Therefore, these demographic or contextual factors are unlikely to have contributed to the observed *Unstuck*-related improvements in flexibility in autistic youth with reliable abstract concept representation.

Importantly, three baseline characteristics appear to be relevant to the observed flexibility improvements following *Unstuck:14-22.* The Stable group was approximately six months younger, about half a standard deviation higher in IQ on average, and had fewer autistic traits across developmental history than the Others group. While our analytic models included these variables as covariates, it is important to consider their potential influence. Younger age and higher IQ in treatment responders is consistent with prior findings from early intensive behavioral and naturalistic developmental interventions (42,43) as well as *Unstuck* (32). Age effects were generally weaker and often became non-significant once adjusted for intellectual functioning (44). Younger children are likely more compliant to study demands consistently throughout the intervention trial, thereby, supporting improved functional outcomes. Higher IQ may facilitate learning intervention-specific vocabulary and scripts and partly reflect stronger fluid and abstract reasoning abilities (45). Lower autistic traits may have facilitated participation in the group-based format of *Unstuck*, which requires sustained interaction and collaboration over the nine-month period. Nevertheless, stable prototype use predicted flexibility improvement after controlling for baseline autistic traits, age, and IQ, suggesting these characteristics were not determining factors in the effectiveness of *Unstuck*.

The present findings have several limitations. First, the Others group represented a heterogeneous category, including individuals with inconsistent representational strategies, unsuccessful learning, or limited engagement. Control analyses comparing the Stable group with only those showing inconsistent strategies yielded a similar pattern of results, although larger samples are needed to confirm this pattern. Larger samples will also be needed to characterize the full range of representational stability, including stable non-prototype strategies, and to examine their associations with treatment response. Second, for individuals with unsuccessful learning or limited engagement, we cannot determine whether this reflected learning and attention difficulties or task demands. Future work should examine whether task adaptations can enhance learning and engagement, helping to clarify whether unsuccessful performance reflects individual learning difficulties or task demands. Finally, the current sample included autistic youth without intellectual disability. Although IQ was higher in the Stable group, IQ alone did not determine task completion or stable prototype use. Replication in samples with a broader range of intellectual abilities will be important for evaluating generalizability.

The present findings have several implications for intervention research in autism. First, prototype-based learning may serve as a promising cognitive marker of intervention responsiveness. Our findings motivate further investigation of its potential as a pre-trial tool for identifying treatment responders and supporting personalized treatment planning. While our longitudinal design included the participant as their own control, it will be important to replicate the current findings in a randomized control design. Furthermore, it is important to examine whether similar predictive profiles can be established over shorter assessment intervals. Second, these finding raise the possibility that prototype-based learning may represent a transdiagnostic cognitive marker of treatment response. Examining whether similar associations emerge across cognitive-behavioral interventions, behavioral targets, and neurodevelopment conditions will help determine broader applicability of this approach. Third, our findings identify abstraction as a potential target for intervention development. *Unstuck:14-22* places strong emphasis on generalizing learned strategies across real-world contexts, aligning with prior intervention studies that have prioritized generalization of treatment effects (46,47). However, few interventions have explicitly targeted the cognitive mechanisms that enable generalization. Future work should examine whether abstraction can be operationalized and scaffolded as an intervention target to strengthen generalization and improve outcomes.

## Conclusion

The present study provides the first evidence for a cognitive mechanistic moderator of responsiveness to cognitive-behavioral treatment for executive function. Together with other moderators of treatment response (12), these findings pave the way for application of approaches for identifying multimodal profiles of treatment response (13,14). Most importantly, our findings illustrate how cognitive strategies measured in the laboratory may shape the learning, transfer, and application of treatment content in everyday life. By bridging basic cognitive science and clinical treatment research, this work opens new avenues for precision treatment in psychiatry.

## Data Availability

All data produced in the present study are available upon reasonable request to the authors

