## Supplementary Materials for "Prototype abstraction predicts response to flexibility intervention in autistic youth"

**Methods**

**Derivation of the final sample**

Out of the 64 participants who completed all assessments at T1, 54 completed the intervention and T3 assessment. Five withdrew before T2, three did not complete the intervention, and an additional two withdrew before T3. For the category learning task, training data was not recorded for two participants due to technical problems (one at T1 and another at T3). In addition, generalization data was not recorded for a different participant at T3 due to technical problems; and another participant did not complete the task at T3.

**Co-occurring conditions and medication**

At enrollment, 41 of 54 participants (76%) were taking at least one psychotropic medication, 26 (48%) were taking at least two, and 11 (20%) were taking at least three. Co-occurring conditions and medication use in the Stable and Others groups are summarized in Table S1.

**Table S1. Co-occurring conditions and medication use at enrollment for Stable and Others groups.**

|  | n (%) | |  |
| --- | --- | --- | --- |
| **Characteristics** | **Stable** | **Others** | Sig. |
| #Participants | 26 | 28 |  |
| **Co-occurring conditions** |  |  |  |
| No co-occurring condition | 1 (4%) | 1 (4%) | p=.99 |
| One co-occurring condition | 4 (15%) | 3 (11%) | p=.70 |
| Two co-occurring conditions | 5 (19%) | 7 (25%) | p=.61 |
| Three or more co-occurring conditions | 11 (62%) | 17 (61%) | p=.18 |
| **Medication** |  |  |  |
| Stimulant ADHD | 16 (62%) | 12 (43%) | p=.17 |
| Non stimulant ADHD | 6 (23%) | 4 (14%) | p=.49 |
| Anti-depressant / Anti-anxiety | 13 (50%) | 19 (68%) | p=.18 |
| Anti-psychotic | 1 (4%) | 4 (14%) | p=.35 |
| Other | 6 (23%) | 9 (32%) | p=.46 |

Categorical variables were compared using the chi-square test when all expected cell counts were greater than 5; otherwise, Fisher's exact test was used.

**Accuracy analysis**

Hierarchical linear modeling (HLM) implemented in MATLAB using the “fitlme” function was used to account for the hierarchical structure of the data. To examine whether Stable and Others groups differed in their acquisition and generalization of category structures across the two pre-intervention time points, two HLMs were fit to examine training and generalization performance. All HLMs shared a common hierarchical structure, in which the measurement units (e.g. training blocks) were nested within timepoint, and timepoint was nested within subjects. Learning effects were represented by a continuous variable coding training blocks (1–5), with positive values indicating learning. Typicality effects were represented by a continuous variable coding feature distance from the prototype (0–3), with negative values indicating a typicality gradient. Each model included the block or typicality effect, timepoint, group (Stable vs. Others), all 2-way interactions and the 3-way interaction, with age, sex assigned at birth (M, F) and IQ as covariates. The three-way interaction was tested initially but was not significant and was therefore removed from subsequent models. Random intercepts and slopes were specified at the subject and subject × timepoint levels. Accuracy for block k, timepoint j, subject i was modeled as:

$$\text{ACC}_{ijk}=\text{intercept}+\beta_{1}\left( \text{block} \right)+\beta_{2}\left( \text{time}\text{point} \right)+\beta_{3}\left( \text{group} \right)+\beta_{43}\left( \text{block}\times\text{time}\text{point} \right)+\beta_{53}\left( \text{block}\times\text{group} \right)+\beta_{6}\left( \text{group}\times\text{time}\text{p}\text{oin}\text{t} \right)+\beta_{\ldots}\left( \text{covariates} \right)+d_{i}+d_{ij}+e_{ijk}$$

(1)

Here, the intercept and β terms are fixed effects, d_i_ is the subject-level random effect, d_ij_ is the measurement-level random effect nested within subjects, and e_ijk_ is the residual. The intercept and β_1_ were allowed to vary across sessions and individuals.

**Results**

**Training performance**

To examine whether learning curves differed between the Stable and Others groups across the two pre-intervention timepoints, we tested block × group, block × timepoint, and timepoint × group interactions. The block × group interaction was significant (t(525)=-2.5, p=.01), indicating different learning curves between groups. Post-hoc analyses showed significant block effects in both groups (Stable: t(249)=10.0, p<.0001; Others: t(274)=4.8, p<.0001), but the Others group exhibited a flatter learning curve than the Stable group (Figure S1A). The timepoint × group interaction was also significant (t(525)=2.3, p=.02). Post-hoc analyses showed that the Stable group showed increased accuracy from T1 to T2 (t(274)=2.8, p=.006), whereas the Others group showed no change (p<.68), suggesting a practice-related improvement in the Stable group only. The main effect of group was not significant (p=.15), indicating no overall difference in training accuracy between groups.

**Generalization performance**

To examine whether typicality effects differed between the Stable and Others groups across the two intervention timepoints, we tested typicality × group, typicality × timepoint, and timepoint × group interactions. Only the typicality × group interaction was significant (t(422)=3.9, p=.0001), indicating different typicality effects between groups. Post-hoc analyses showed significant typicality effects in both groups (Stable: t(202)=-14.9, p<.0001; Others: t(218)=-5.1, p<.0001), but the effect was weaker in the Others group (Figure S1B). The main effect of group was significant (t(422)=-4.5, p<.0001), indicating that the Stable group showed overall higher generalization accuracy. The main effect of timepoint was not significant (p=.68).

**Familiarity effect in generalization**

To examine whether familiarity effect, defined as the difference in accuracy between training and novel 2-away items, differed between the Stable and Others groups across the two intervention timepoints, we tested familiarity × group, familiarity × timepoint, and timepoint × group interactions. None of the interactions were significant, as well as the main effect of familiar (ps≥.06), suggesting that familiarity for individual exemplars did not aid categorization performance. As for the main effect of group, better performance in the Stable group was expected (t(206)=-4.3, p<.0001).

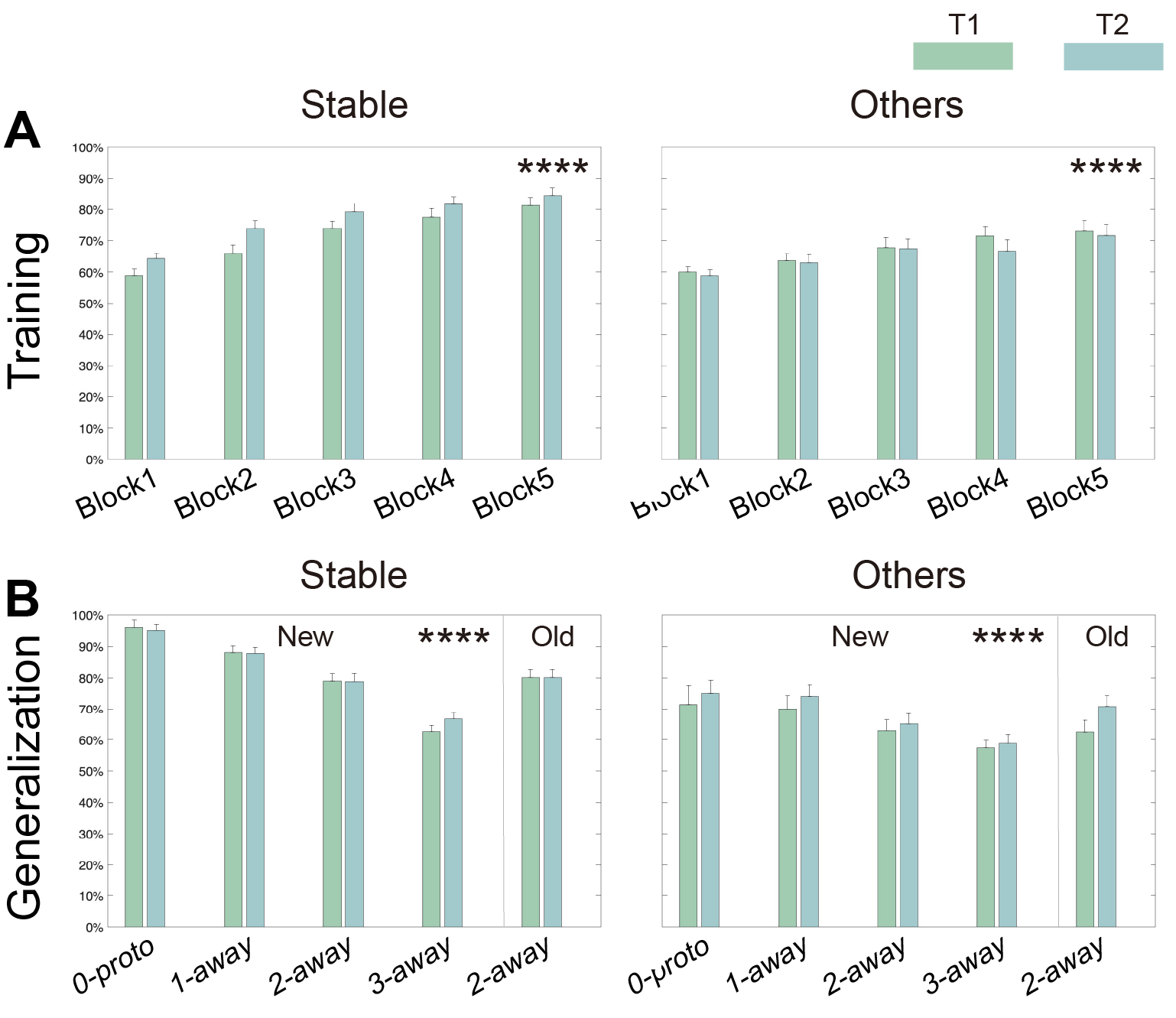

**Figure S1 Training and generalization performance in the Stable and Others groups**

(A) Training accuracy (mean % correct) by block for each timepoint. (B) Generalization accuracy for new stimuli at each feature distance (0, 1, 2, 3) and for old (training) items at 2-away for each time-point. Asterisks indicate significant learning effects in A and typicality effects in B (p < .0001, hierarchical linear modeling). Error bars represent standard error of the mean.

**Appendix 1 Consort Diagram**

COMPLETED: Decline/Excluded Total (n=144)

Declined to participate (n=83)

- Logistics (Locations, time commitment, etc) (n=49)
- Decision not to participate in group therapy (Teen declined) (n=17)
- Lost to follow up (n=17)

Excluded (n=61)

- Unable to scan (n= 39)
- Unable to partake in group (dx, IQ, language barriers, age requirements) (n= 21)
- Previously completed novel task (pilot) (n=1)

Assessed for Eligibility (n= 208)

Enrolled in Study & Completed both category learning task in MRI and behavioral assessments at TP1 baseline (n=64)

COMPLETED: Withdrew/Excluded Total (n=5)

Withdrew (n=4)

- Reason:
  - LM12 – no longer interested
  - LM35 – logistics for assigned group
  - LM54 – moved (out of country)
  - LM61 – new dx and needed more time to process before doing therapy.

Excluded (n=1)

- Reason:
  - LM24 – clinically not a good match for group

Completed both category learning task in MRI and behavioral assessments at TP2 pre intervention (n=59)

COMPLETED: Withdrew/Excluded Total (n=5)

Withdrew (n=2)

- Reason:
  - LM37 – declined to consent at age of majority
  - LM65 – felt like group wasn’t helpful

Discontinued intervention, but remained in research (n=3)

- Reason
  - LM11 – Not good fit for group (Excluded)
  - LM21 – parents felt that group was not helpful (WD)
  - LM02 – Driving distance from group (WD)

Completed intervention

(n=54)

COMPLETED: Withdrew/Excluded Total (n=1)

TP3 includes 3 participants who discontinued the intervention early, but still participated in research appointments.

Excluded (n=1)

- LM38 – Participants had clinically significant cysts on MRI.

Completed both category learning task in MRI and behavioral assessments at TP3 endpoint (n=56)

Analysis for the present study (n=54)
